# Reversion of the inflammatory markers in patients with chronic limb threatening ischaemia

**DOI:** 10.1101/2023.07.25.23293181

**Authors:** Joana Ferreira, Susana Roque, Alexandre Lima Carneiro, Adhemar Longatto-Filho, Isabel Vila, Cristina Cunha, Cristina Silva, Amílcar Mesquita, Jorge Cotter, Margarida Correia-Neves, Armando Mansilha, Pedro Cunha

**Author notes:** Joana Ferreira, Centro Hospitalar Universitário de São João, 4200-319, Porto, Portugal.

## Abstract

**Introduction:** Peripheral arterial disease (PAD) affects over 202 million individuals worldwide, with 1.3% suffering from chronic limb-threatening ischaemia (CLTI). Atherosclerosis, characterized by intense inflammation, is the primary cause of PAD. Inflammation is linked to higher mortality rates, especially in CLTI patients. This study aims to compare the evolution of inflammatory markers between claudication and CLTI patients over three, six, and twelve months.

**Methods:** A prospective, single-center observational study was conducted from January 2018 to July 2022. A PAD cohort was observed at admission and at three, six, and twelve months, with data on clinical, analytical, and inflammatory markers collected. The analyzed markers included positive acute phase proteins (C-reactive protein - CRP and fibrinogen) and negative acute phase proteins (albumin, total cholesterol, and high-density lipoprotein - HDL).

**Results:** The study involved 119 subjects (mean age: 67.58 ± 9.60 years; 79.80% males), with 65 patients having claudication and 54 with CLTI. At admission, CLTI patients exhibited significantly higher serum levels of CRP and fibrinogen (positive acute phase proteins) and lower levels of albumin, total cholesterol, and HDL (negative acute phase proteins) compared to claudication patients. Three months after CLTI resolution, negative acute phase proteins increased, and positive acute phase proteins decreased. However, no significant changes in inflammatory proteins were observed in patients with claudication over time.

**Conclusion:** CLTI patients demonstrate an inflammatory state, which may have deleterious consequences and be partially reversible after the resolution of the ischemic process. Recognizing the potential for reversibility through revascularization/amputation underscores the significance of timely intervention.

## 1. INTRODUCTION

Lower extremity peripheral arterial disease (PAD) is characterized by atherosclerosis occurring in the lower limbs and affects over 230 million adults worldwide[1,2]. Due to its high prevalence in low and middle-income countries PAD is becoming an increasingly global problem. The most advanced stage of PAD is chronic limb threatening ischemia (CLTI) which accounts for 11% of diagnosed PAD cases[1]. Patients with CLTI have a worse prognosis, with at least 20% of them dying within one year of diagnosis[3].

PAD and inflammation are particularly connected. Inflammation plays a significant role in the initiation and progression of atherosclerosis, particularly in PAD[4,5]. Traditional cardiovascular risk factor exerts their proatherogenic role, at least in part, through an inflammatory mechanism[4,6]. Also important is to acknowledge the important role of inflammation on the acceleration of the systemic arteriosclerotic process, leading equally to terminal cardiovascular and renal disease[7,8]. Pro-inflammatory cytokines [interleukin-6 (IL-6) and tumor-necrosis factor-α (TNFα), TNF soluble receptor-II (TNFαSRII)] are strongly linked to PAD[6]. The lower limbs, which possess a large vascular bed, frequently harbor inflamed plaques that release inflammatory mediators. These mediators contribute to the development of coronary artery disease, thereby explaining the higher incidence of coronary artery disease in PAD patients[4]. The prevalence of coronary artery disease in PAD ranges from 43% to 90%, while the prevalence of PAD in coronary artery disease patients is less than 25%[4]. Studies have shown that the severity of coronary atherosclerosis is related with the degree of inflammatory response in the affected PAD limb[4]. Consequently, it has been suggested that it is not PAD itself, but rather its systemic inflammatory activity, that is associated with an increased number of coronary events[4].

The main aim of this study is to determine the evolution of the inflammatory parameters at three, six and twelve months in patients with claudication and with CLTI.

The second objective is to assess the differences in inflammatory parameters between patients with claudication and those with CLTI, through time.

## 2. METHODS

### 2.1. Study Type, Inclusion/Exclusion Criteria, Ethical Considerations

An observational, prospective, study was conducted from January 2018 to July 2020 at a single institution. Consecutive patients with PAD, attending the Vascular Surgery consultations or admitted at the Vascular Surgery ward who fulfilled the required criteria were included. Inclusion criteria: Patients with PAD suggested by the clinical history and objective examination and, confirmed with ankle-brachial index (ABI). Exclusion Criteria: Any disease responsible for body composition changes or pro-inflammatory state in the last three months. Ethics approval was obtained from the local Hospital, with the protocol number 75/2017. All the participants signed the informed consent.

### 2.2. Clinical Characteristics

Patient’s age, gender, PAD clinical stage (CLTI and claudication), arterial hypertension, diabetes, dyslipidemia, smoking habits and medication were collected at admission and defined as previously stated [9]. Fontaine stage III was defined as persistent rest pain for more than two weeks [10]. Fontaine stage IV was defined as ischemic skin lesions [10]. Both cases were confirmed with the following hemodynamic parameters: ankle pressure below 50mmHg, or absent palpable ankle pulses or toe pressure below 30mmHg in patients with diabetes and incompressible vessels [11].

The patients were schedule for an evaluation at three, six and twelve months. The clinical evolution, amputation rate (major and minor), death and causes of death were registered.

### 2.3. Inflammatory Parameter

The serum inflammatory parameters determined were positive acute phase proteins (C-reactive Protein-CRP- and fibrinogen) and negative acute phase proteins (albumin, total cholesterol and high-density lipoprotein-HDL).

Blood Samples were collected after a 10–12 hours fast in the morning, taken into appropriate Vacutainer, centrifuged within 5min for 4000 cycle/minute, and serum was separated. The serum inflammatory parameters were evaluated at admission, three, six and twelve months of follow up, and tests were performed by routine procedures in the department of clinical chemistry.

### 2.4. Statistical Analysis

Continuous variables were expressed as the mean ± standard deviation (SD) and as the percentage for categorical variables. The Shapiro–Wilk test was used to assess all continuous variables for normality. Between-group differences in continuous variables were assessed with Student’s t-test or with the Mann-Whitney U test, and the effect size r was determined by calculating the ratio between test statistics Z and the square root of the number of pairs n. Categorical variables between two groups were compared with Chi-square. Wilcoxon Signed Rank test and paired samples t-test were used to study the evolution of acute phase proteins and analytic data, comparing 4 time points (t = admission; t = 3 months, t = 6 months and t = 12 months).

A p-value of less than .05 was considered significant. Statistical evaluation was performed using SPSS software, version 20.0 (SPSS, Inc., Chicago, IL, USA).

## 3. RESULTS

### 3.1. Clinical Characteristics at Admission

A total 119 patients (95 men) with an average age of 67.58 ± 9.60 years-old with PAD were enrolled in the study. The group of patients was evenly distributed as 65 patients (54.62%) had claudication and 54 (45.38%) had CLTI. Nineteen of these patients had rest pain.

No differences were registered between patients with claudication and CLTI on age, gender, cardiovascular risk factors and medication, except on smoking habits (Table I). There was a significant higher prevalence of smokers and a higher smoking load in claudication group [Claudication: Median = 40.00; IQR = 45.00; CLTI: Median = 13.00; IQR = 40.00; p = 0.031] (Table 1).

**Tab. 1.** Cardiovascular risk factor and comorbidities of the PAD population.

|  | **Claudication (n=65)** | **CLTI (n=54)** | **$\chi^2$** | **df** | **p-value** | **Phi** |
| --- | --- | --- | --- | --- | --- | --- |
| **Male (n; %)** | 55; 84.61 | 40; 74.07 | 1.555 | 1 | 0.212 | 0.115 |
| **Hypertension (n; %)** | 49; 75.38 | 37; 68.52 | 0.694 | 1 | 0.405 | 0.076 |
| **Dyslipidemia (n; %)** | 41; 63.08 | 32; 59.26 | 0.181 | 1 | 0.670 | 0.039 |
| **Smoker/ex-smoker (n; %)** | 53; 81.53 | 30; 55.56 | 7.969 | 1 | **0.005\*** | 0.261 |
| **Diabetes (n; %)** | 23; 35.38 | 28; 51.85 | 3.266 | 1 | 0.071 | - 0.166 |
| **Coronary artery disease (n; %)** | 9; 13.85 | 6; 11.11 | 0.200 | 1 | 0.654 | 0.041 |
| **Statins (n;%)** | 58; 89.23 | 44; 81.48 | 0.961 | 1 | 0.327 | 0.090 |
| **Fibrate (n;%)** | 4; 6.15 | 5; 9.26 | Fisher | - | 0.512 | 0.063 |
| **Ezetimibe (n;%)** | 3; 4.615 | 3; 5.55 | Fisher | - | 0.812 | 0.022 |
| **Antiplatelet (n;%)** | 54; 83.08 | 40; 74.07 | 1.042 | 1 | 0.307 | 0.094 |
| **ACEi/ARA (n;%)** | 20; 30.77 | 16; 29.63 | 0.000 | 1 | 0.989 | 0.001 |
| **Beta-blockers (n;%)** | 13; 20.00 | 11; 20.37 | 0.003 | 1 | 0.953 | 0.005 |

**Note:** Application of Chi-Square test and Fisher's exact test.
**ACEi:** angiotensin-converting enzyme inhibitors.
**ARA:** angiotensin receptor antagonists
**CLTI:** chronic limb-threatening ischemia

### 3.2. Clinical Evolution

During the follow-up, 28 (23.53%) patients had completed four clinical visits and reached the end of the follow-up; 52 (43.70%) patients had three appointments and 93 (78.15%) two. Out of the 65 patients with claudication, 14 (21.54%) improved their symptoms with medical therapy, five (7.69%) had worsening symptoms and one patient was readmitted due to CLTI. Six (5.04%) deaths were registered: five in patients with CLTI, one in a patient with claudication. The causes of death were: acute myocardial infarction (3 patients); heart failure (2 patients); pulmonary infection (1 patients); lung cancer (1 patient). Thirty patients (55.56%) with CLTI were submitted to minor amputation [23 (42.59%) and 12 (22.22%) to major amputations].

### 3.3. Inflammatory Parameters

At admission patients with CLTI had significantly higher serum levels of positive acute phase proteins (CRP, and fibrinogen) and lower serum level of negative acute phase proteins (albumin, total cholesterol and HDL), when compared to patients with claudication (Table 2).

**Tab. 2.**
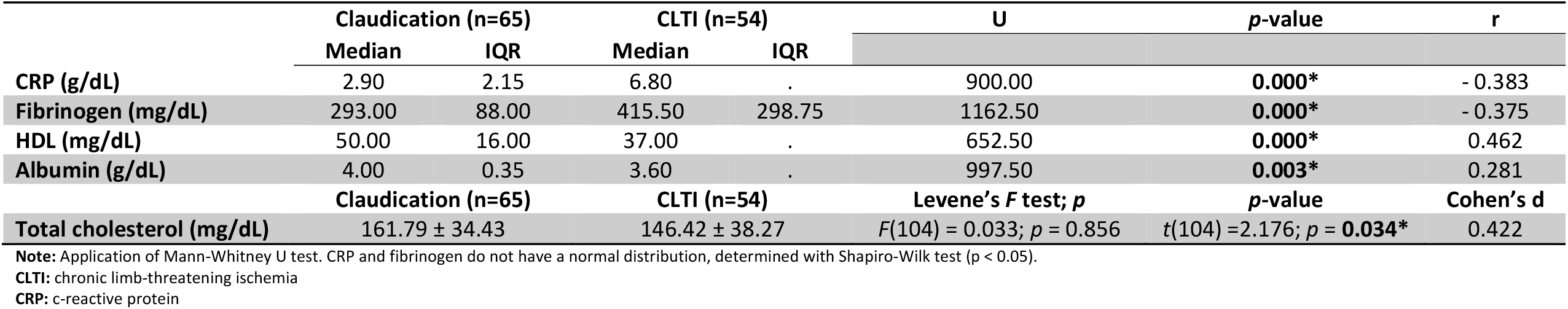
Positive (CRP and fibrinogen) and negative (HDL, albumin and total cholesterol) acute phase proteins determined in PAD patients, at admission.

No differences were found in patients with CLTI, between Fontaine stage III and IV on the serum level of acute phase proteins (Table 3).

**Tab. 3.**
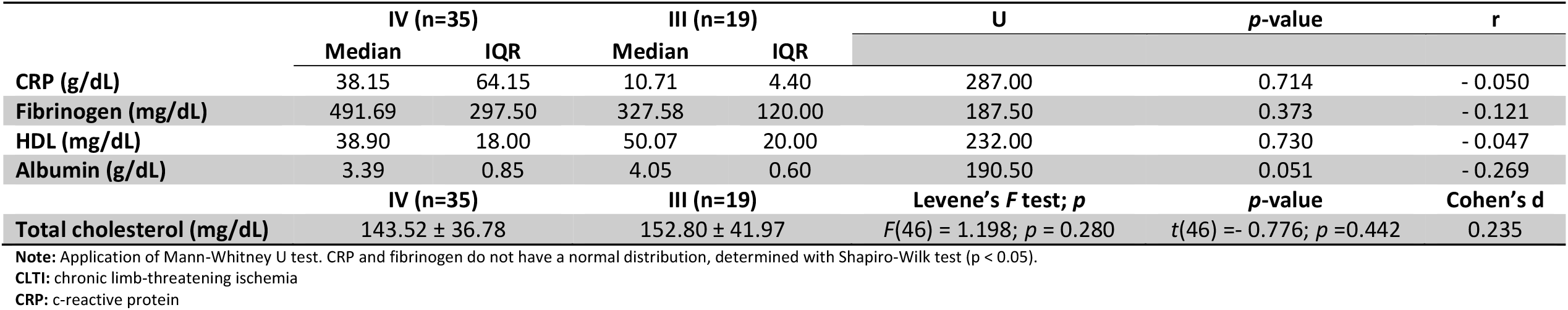
Positive (CRP and fibrinogen) and negative (HDL, albumin and total cholesterol) acute phase proteins determined in patients with CLTI (Fontaine III vs Fontaine IV), at admission.

### 3.4. Evolution of Inflammatory Parameters

Analyzing the evolution of acute phase proteins patients, we noted that three months after the resolution of the ischemic process in patients with CLTI (by amputation or revascularization) there was an increase in serum levels of HDL and albumin (negative acute phase proteins) and decrease in CRP and fibrinogen (positive acute phase proteins (Table 4). These differences were not registered at six or twelve months (Table 4). No differences were noted through time in patients with claudication (Table 5)

**Tab. 4.**
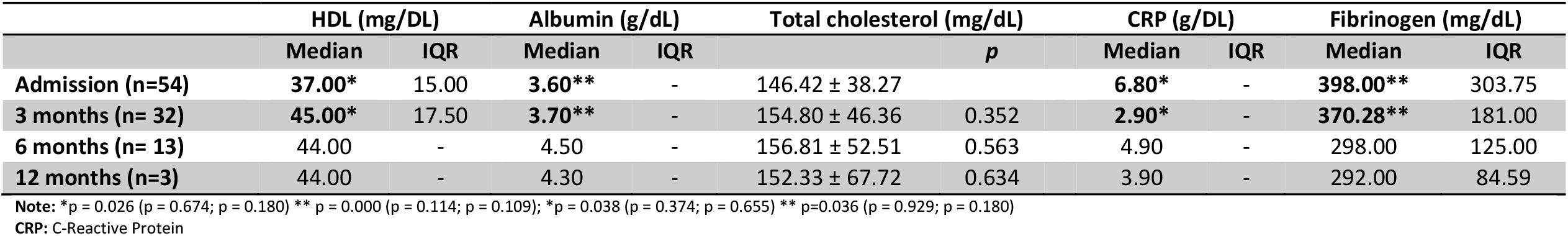
Evolution of negative acute phase proteins (HDL, albumin and total cholesterol) and positive acute phase proteins (CRP and fibrinogen) in patients with CLTI at three, six and twelve months.

|  | HDL (mg/DL) |  | Albumin (g/dL) |  | Total cholesterol (mg/dL) | *p* | CRP (g/DL) |  | Fibrinogen (mg/dL) |  |
| --- | --- | --- | --- | --- | --- | --- | --- | --- | --- | --- |
|  | Median | IQR | Median | IQR |  |  | Median | IQR | Median | IQR |
| **Admission (n=54)** | **37.00\*** | 15.00 | **3.60\*\*** | - | 146.42 ± 38.27 |  | **6.80\*** | - | **398.00\*\*** | 303.75 |
| **3 months (n= 32)** | **45.00\*** | 17.50 | **3.70\*\*** | - | 154.80 ± 46.36 | 0.352 | **2.90\*** | - | **370.28\*\*** | 181.00 |
| **6 months (n= 13)** | 44.00 | - | 4.50 | - | 156.81 ± 52.51 | 0.563 | 4.90 | - | 298.00 | 125.00 |
| **12 months (n=3)** | 44.00 | - | 4.30 | - | 152.33 ± 67.72 | 0.634 | 3.90 | - | 292.00 | 84.59 |

**Note:** \**p* = 0.026 (*p* = 0.674; *p* = 0.180) \*\* *p* = 0.000 (*p* = 0.114; *p* = 0.109); \**p* = 0.038 (*p* = 0.374; *p* = 0.655) \*\* *p*=0.036 (*p* = 0.929; *p* = 0.180)
**CRP:** C-Reactive Protein

**Tab. 5.** Evolution of negative acute phase proteins (HDL and albumin) and positive acute phase proteins (CRP and fibrinogen) in patients with claudication at three, six and twelve months.

|  | HDL (mg/DL) |  | Albumin (g/dL) |  | CRP (g/DL) |  | Fibrinogen (mg/dL) |  |
| --- | --- | --- | --- | --- | --- | --- | --- | --- |
|  | Median | IQR | Median | IQR | Median | IQR | Median | IQR |
| **Admission (n=65)** | 50.00 | 16.00 | 4.00 | 0.35 | 2.90 | 2.15 | 293.00 | 88.00 |
| **3 months (n=61)** | 53.00 | 19.00 | 3.90 | 0.32 | 3.10 | 2.33 | 311.00 | 90.00 |
| **6 months (n=39)** | 53.00 | 54.00 | 3.95 | 0.33 | 3.70 | 6.33 | 295.00 | 143.00 |
| **12 months (n=25)** | 49.00 | 17.00 | 4.10 | 0.73 | 2.90 | 1.42 | 294.00 | 96.50 |

**Note:** Application of Friedman test. HDL and albumin do not have a normal distribution, determined with Shapiro-Wilk test (\*< 0.05).
**CRP:** c-reactive protein

We also noted that for most inflammatory markers the difference between those who had claudication and patients with CLTI was present at three months but disappeared at six and twelve months. (Fig. 1)

**Fig. 1.**
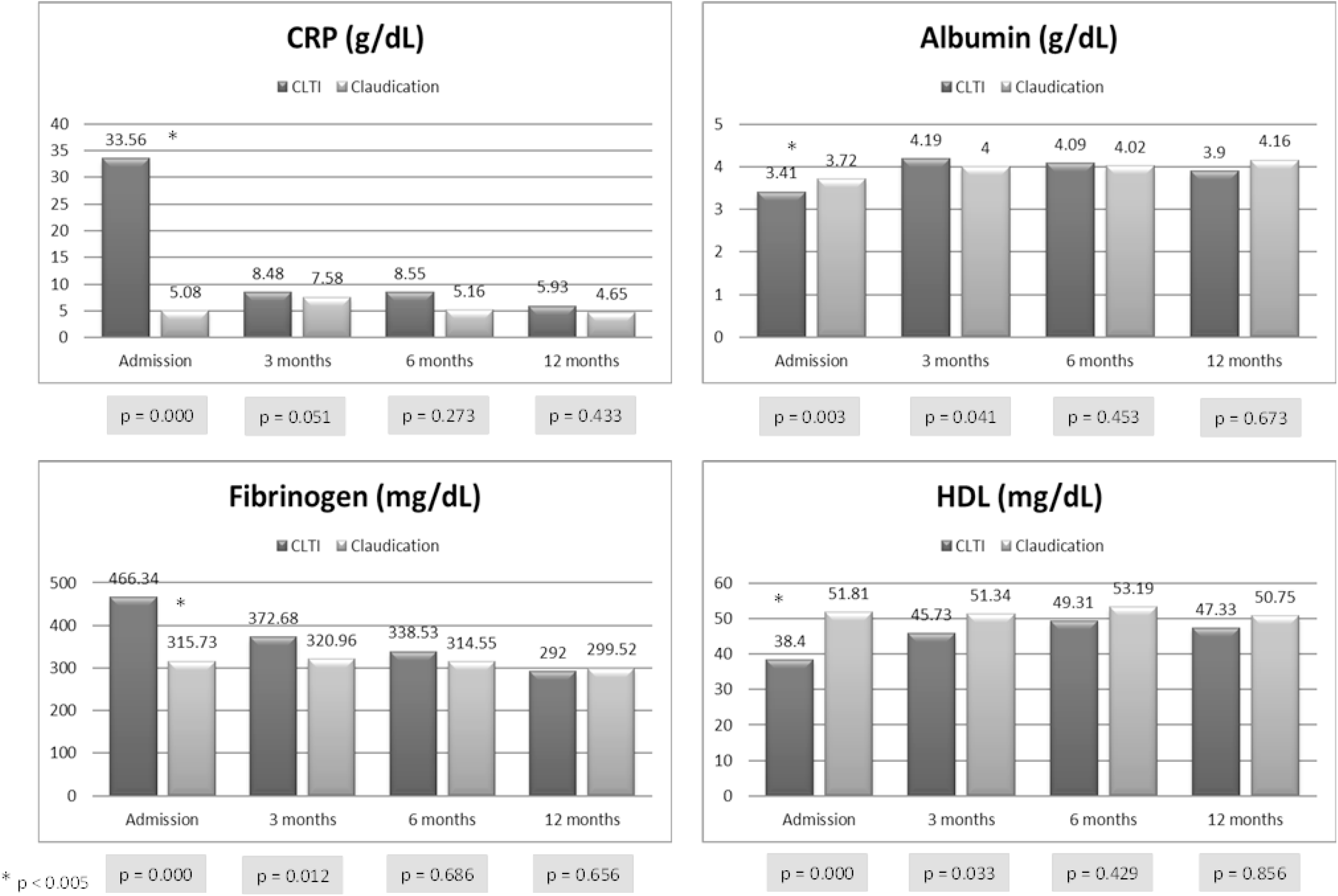
Comparison of acute phase proteins between patients with claudication and with CLTI at three, six and twelve months.

## 4. DISCUSSION

To the best of our knowledge, this paper is one of the first reporting the possibility of partially reversing the inflammatory state in patients with CLTI, after resolution of the critical limb ischemia event. The novelty of this study lies in the observation that three months following the resolution of the ischemic process, there is a reversal of the inflammatory markers, and the differences between CLTI and claudicants disappear at six months. To our knowledge, no other study has compared the inflammatory markers between CLTI and claudicants over such an extended period of time.

### 4.1. Inflammatory Parameters in CLTI and Patients with Claudication

We observed that patients with CLTI have a significant higher level in CRP and fibrinogen (positive acute phase proteins) and a significant lower serum level of albumin, total cholesterol and HDL (negative acute phase proteins) compared to patients with claudication, as we have previously demonstrated [9].

We hypothesized that this inflammatory pattern could be caused by the extension of atherosclerosis lesions, tissue infection or the ischemic tissue process. It has been reported that the severe reduction in tissue oxygenation present in patients with CLTI, can trigger an inflammatory reaction, leading to an increase in fibrinogen levels [12]. The elevated fibrinogen levels result in higher blood viscosity, promoting thrombosis, and potentially explaining the higher risk of graft occlusion, stroke, myocardial infarction or death observed in CLTI patients [3,12]. Increased plasma viscosity also impairs the vessel perfusion, particularly in small vessels such as capillaries, increasing tissue ischemia [13].

The inflammatory reaction also contributes to the generation of a proatherogenic lipid profile characterized by low HDL, oxidized LDL and high triglycerides [14]. We observed that the CLTI patients had low HDL and high triglyceride levels. Additionally, patients with CLTI had a lower albumin level which are considered a powerful predictor of mortality [14].

To investigate whether the inflammation in CLTI patients is primarily caused by tissue infection, we compared the serum levels of acute phase proteins between patients in Fontaine stage III and stage IV, but no differences were found, as suggested by other authors [13].

Our hypothesis was that the differences in inflammatory markers between CLTI and claudicants could be explained by the activity and extension of the atherosclerotic process in patients with critical limb ischemia. However, the pattern of evolution of the inflammatory markers (that we discuss below) suggests that it is the ischemic tissue process, rather than the extension of atherosclerotic plaques, that causes the differences between CLTI and claudicants.

### 4.2. Evolution of Inflammatory Parameters

We observed that three months after the resolution of the CLTI state (by amputation or surgery), there was a reversion, at least in part, of the inflammatory parameters, with an increase in the negative acute phase proteins (HDL and albumin) and a decrease in positive acute phase proteins (CRP and fibrinogen). These differences were not observed in patients with claudication or at other periods of time in patients with CLTI. At three months, patients with CLTI maintained a higher serum level of CRP and fibrinogen and lower levels of albumin compared to patients with claudication. However, these differences disappeared at six months. Our findings align with two previous studies: Bismuth et al. reported a decrease in CRP and an increase in HDL, and albumin after revascularization in 30 patients with CLTI [15]. Woodburn et al. compared fibrinogen levels in 56 patients with CLTI at admission and 16 weeks after revascularization, observing a reduction in fibrinogen levels, although they remained higher than those of the controls [3]. The normalization of fibrinogen levels following successful vascular surgery of critically ischemic limbs (revascularization or amputation) suggests that tissue ischemia may stimulate hepatic fibrinogen synthesis, possibly through interleukin-6 produced by activated monocytes [12]. Both of these studies analyzed the data at three months, and no other study has examined the data over a longer follow-up period.

### 4.3. Clinical Implications

Demonstrating that following the resolution of CLTI that there is a decrease in serum levels of inflammatory markers suggests that the deleterious impact of inflammation can be mitigated with a prompt intervention.

This study also highlights the potential role of anti-inflammatory medical therapy in PAD patients. For example, cankinumab is an interleukin-1β inhibitor that improved walking performance in patients with claudication while reducing blood CRP and IL-6 [16]. Statins have been associated with a reduction in systemic inflammation, as measured by CRP, and have shown benefits in terms of lower mortality, reduced major adverse cardiovascular and cerebrovascular events, and longer amputation-free survival in CLI patients [6,16,17]. The benefit of statins are closely related to their anti-inflammatory effect [18]. Fibrates also exhibit fibrinogen-lowering action[6].

### 4.4. Strengths and Limitations

Strengths of the paper:

1. Comprehensive investigation: The paper explores the association between PAD and inflammation, and the evolution of inflammatory parameters in patients with claudication and those with CLTI over a period of three, six, and twelve months.
2. Comparison between claudication and CLTI patients over an extended period of time: The study compares patients with claudication and CLTI in terms of their inflammatory parameters. By examining the differences between these two groups, the researchers can better understand the impact of disease severity on inflammation.
3. Findings on the reversal of inflammatory state in CLTI patients: The study reports that three months after the resolution of the ischemic process, there is a partial reversal of the inflammatory state in CLTI patients. This finding suggests that prompt intervention and anti-inflammatory therapies may help alleviate the detrimental effects of inflammation in PAD patients.
4. Comparing the Stage III and IV of Fontaine Classification: The paper reports no difference in the serum levels of acute phase proteins between patients on stage III and IV. This fact underlines the role of the ischemic tissue in the inflammatory changes registered in patients with CLTI.

This research work as several limitations:

1. Sample size: The study included a total of 119 patients, followed by the main author which may not be representative of the entire PAD population followed at our institution. A larger sample size could provide more robust and generalizable results.
2. Prevalence of men in the studied population: Approximately 80% of the recruited sample were men. As such the results cannot be generalized to all population.
3. Short follow-up period (even if the longest recorded in the literature): The study evaluated the evolution of inflammatory parameters at three, six, and twelve months. However, a longer follow-up period could provide more insights into the long-term changes and trends in inflammatory markers. However, as far as we know this is the longest follow-up published.
4. Single-center study: The study was conducted at a single institution, which may limit the generalizability of the findings. A multi-center study could provide a more comprehensive understanding of the topic.
5. Other factors: The paper did not investigate other potential factors that could influence the inflammatory state, as the genetic, the practice of exercise or dietary habits.
6. Causality: This research work is also not able to establish any a causal relationship between the resolution of the ischemia process and the improve in the inflammatory data.

## 5. CONCLUSIONS

This study provides valuable insights into the evolution of inflammatory parameters in patients with PAD, specifically comparing those with CLTI to claudicants. It emphasizes that patients with CLTI exhibits an inflammatory state that can be at least partially reverted after the resolution of the ischemic process, thereby mitigating the negative impact of inflammation.

## Data Availability

In case data is requested, it will be provided.

## CONFLITS OF INTEREST STATEMENT

The authors have no conflicts of interest to declare.

## AUTHOR’S CONTRIBUTIONS

Joana Ferreira – Conceived of the presented idea; wrote the manuscript, collected and analysed the data. Susana Roque - Determined the serum level of inflammatory markers, conceived the methods relating to the laboratory measurements of myokines, revised the manuscript. Alexandre Lima Carneiro – Helped in the manuscript writing, in the data analysis and made the statistical analysis. Adhemar Longatto-Filho-Analyzed the data and revised the manuscript. Isabel Vila – Collected the clinical data, helped in the manuscript writing and revised the paper. Cristina Cunha - Collected the clinical data, helped in the manuscript writing and revised the paper. Cristina Silva - Collected the clinical data, helped in the manuscript writing and revised the paper. Amílcar Mesquita – Revised the paper and contributed to the final manuscript. Jorge Cotter – Revised the paper and contributed to the final manuscript. Margarida Correia-Neves – Revised the paper and contributed to the final manuscript. Armando Mansilha – Supervise the paper, revised the paper and contributed to the final manuscript. Pedro Cunha – Contributed to the design, collected the data, revised the paper and contributed to the final manuscript.

